# A multiscale characterization of cortical shape asymmetries in early psychosis

**DOI:** 10.1101/2023.04.29.23289297

**Authors:** Yu-Chi Chen, Jeggan Tiego, Ashlea Segal, Sidhant Chopra, Alexander Holmes, Chao Suo, James C. Pang, Alex Fornito, Kevin M. Aquino

**Author notes:** Correspondence to: Yu-Chi Chen, Brain and Mind Centre, University of Sydney, 100 Mallett Street, New South Wales, 2050, Australia. These authors contributed equally to this work (James C. Pang, Alex Fornito, Kevin M. Aquino).

## Abstract

**Background:** Psychosis has often been linked to abnormal cortical asymmetry, but prior results have been inconsistent. Here, we applied a novel spectral shape analysis to characterize cortical shape asymmetries in patients with early psychosis across different spatial scales.

**Methods:** We used the Human Connectome Project for Early Psychosis dataset (aged 16–35), including 56 healthy controls (male = 37, female = 19) and 112 patients with early psychosis (male = 68, female = 44). We quantified shape variations of each hemisphere over different spatial frequencies and applied a generalized linear model to compare differences between healthy control participants and patients with early psychosis. We further used a canonical correlation analysis (CCA) to examine associations between shape asymmetries and clinical symptoms.

**Results:** Cortical shape asymmetries, spanning wavelengths between about 22 mm and 75 mm, were significantly different between healthy control participants and patients with early psychosis (Cohen’s *d* = 0.28−0.51), with patients showing greater asymmetry in cortical shape than controls. A single canonical mode linked the asymmetry measures to symptoms (CCA *r* = 0.45), such that higher cortical asymmetry was correlated with more severe excitement symptoms and less severe emotional distress. In contrast, significant group differences in morphological asymmetries of cortical thickness, surface area, and gyrification at either global or regional levels were not identified.

**Conclusions:** Cortical shape asymmetries are more sensitive than other morphological asymmetries in capturing abnormalities in patients with early psychosis. These abnormalities are expressed at coarse spatial scales and are correlated with specific symptom domains.

**Highlights:**

- Cortical shape asymmetries are more sensitive than other cortical asymmetry measures, such as cortical thickness, surface area and gyrification, in capturing abnormalities in patients with early psychosis.
- The abnormalities in cortical shape asymmetry are expressed at coarse spatial scales and are correlated with excitement and emotional distress symptoms.

## 1. Introduction

Relative to other primates, the human cerebral cortex shows a greater degree of anatomical and functional asymmetry between the left and right hemispheres and greater inter-individual variability in this asymmetry (1–3). Accordingly, hemispheric asymmetries have been implicated in the evolution of human-specific cognition and behavior (4, 5). Conversely, abnormal hemispheric lateralization has been linked to psychosis (6–8), with some proposing that it is linked to the unique expression of this syndrome in humans (9). However, reported links between abnormal asymmetry and psychosis have not been consistently replicated (7, 10–14).

Brain asymmetries are commonly considered on a population basis by mapping average asymmetry levels across a group of individuals (4, 11, 15–17), which is also referred to as directional asymmetry (2, 18). For instance, the left planum temporale, encompassing Wernicke’s area, is substantially larger, on average, than its right-hemisphere counterpart in healthy control (HC) participants (5). Interestingly, patients with schizophrenia and their relatives show reduced asymmetry in planum temporale compared to HCs (8, 19, 20). However, population-based studies are not entirely informative as cortical asymmetry is highly variable between individuals (2, 3, 18, 21, 22), with many people showing little or even reversed asymmetries relative to the population average (2, 3, 18, 23). Individual deviations of asymmetry from the population mean are referred to as fluctuating asymmetries and may be driven by environmental stress, developmental instability/plasticity, or individual-specific genetic perturbations (18, 22, 24–27). Studies of fluctuating asymmetries in psychosis are scarce, but a study by Núñez et al. (28) has shown that such asymmetries in cortical shape at the global level are increased in patients with schizophrenia and are associated with negative symptoms.

One factor that complicates the identification of reliable asymmetry phenotypes in psychosis is that most analyses to date have focused either on defined regions-of-interest (ROI) or have relied on point-wise (e.g., voxel-based) analyses, which only consider asymmetries at certain spatial resolution scales (19, 20, 29, 30). However, cortical asymmetries can be identified at multiple scales, ranging from entire hemispheres (e.g., Yakolevian torque (31)) to more fine-scale sulcal and gyral features (32). Whether abnormal asymmetries in psychosis are expressed at certain specific scales or are a multiscale phenomenon remains unclear. Moreover, most anatomical asymmetry studies use size-related measures, such as volume, cortical thickness, and surface area, which often conflate individual differences in size and shape and have limited sensitivity for capturing individually unique properties of brain anatomy (21, 33).

We have recently shown that multiscale spectral descriptions of asymmetries in cortical shape, rather than size, are highly personalized, akin to a cortical fingerprint, and can identify subjects more accurately than common morphological measures (e.g., volume, cortical thickness, and surface area), measures of inter-regional functional coupling, and the cortical shapes of individual hemispheres (21). Mathematically, these spectral shape descriptions are obtained through eigen-decomposition of the Laplace-Beltrami Operator (LBO) of the cortical surface (21, 34, 35). The resulting eigenfunctions correspond to an orthogonal basis set of spatial patterns of different spatial frequencies that capture the geometry of the cortex, and the eigenvalues represent their spatial frequencies (see Methods) (33–35). Analysis of these spatial eigenfunctions is ubiquitous in many branches of physics, engineering, and biology (33–35) and naturally captures geometric properties from the coarse scale (low-order eigenfunctions) to the fine scale (high-order eigenfunctions) forming a multiscale description of geometry (21, 34). This multiscale description departs from conventional methods focusing on specific regions or global hemispheric differences (10, 15, 19, 20, 30, 36, 37). Our previous work has shown that optimal subject identifiability of cortical shape asymmetry occurs at coarse spatial scales, corresponding to wavelengths larger than about 37 mm. We also found that these coarse scale asymmetries are correlated with individual differences in general cognitive function and that they are largely driven by unique environmental, rather than genetic, factors (21). Together, these findings suggest that spectral shape analysis of cortical asymmetries offers a window into understanding highly personalized features of brain anatomy.

Here, we applied spectral shape analysis to investigate cortical asymmetries in patients with early psychosis (EP), who were within five years of the initial psychosis onset. EP is a key period to understand brain changes associated with the development of psychosis that is less confounded by prolonged treatment exposure (38). Prior works have shown abnormal cortical asymmetry in patients with psychosis (19, 20, 28, 29, 36), and cortical shape asymmetry at coarse spatial scales captures the most individualized and robust information (21). Thus, we aim to examine the scale-specific cortical shape asymmetry on EP patients and HCs. Moreover, the multiscale shape descriptions isolate scale-specific and shape-specific effects that traditional methods cannot identify. We further compared the cortical shape asymmetry with asymmetries based on cortical thickness, surface area, and local gyrification index (LGI). Finally, we explored the relationships between cortical shape asymmetry and different psychotic symptoms.

## 2. Methods

### 2.1. Neuroimaging data

### 2.1.1. Human Connectome Project for Early Psychosis (HCP-EP)

We used open-source data from the Human Connectome Project for Early Psychosis (HCP-EP; https://www.humanconnectome.org/study/human-connectome-project-for-early-psychosis; (38)), which includes 169 subjects with preprocessed structural MRI data. We excluded data for one individual who showed eigen-groups of matched asymmetry signature (MAS) values that were more than three standard deviations below the sample mean. The remaining 168 participants include 112 patients with early psychosis (EP; aged 16–34, mean = 23.26, standard deviation = 3.67; male = 68, female = 44) and 56 healthy controls (HCs; aged 16–35, mean = 24.10, standard deviation = 4.47; male = 37, female = 19). For EP patients, the Chlorpromazine Equivalence (CPZ) of current antipsychotic drug was between 0 to 1000 mg/d (mean = 165 mg/d), and the exposure time to antipsychotic medication was between 0 to 56 months (mean = 14.3). The patients were diagnosed with non-affective (n = 80) and affective (n = 32) psychosis and were all within five years of the initial onset of their psychotic symptoms. The criteria for non-affective psychotic disorders in this study are the diagnosis of schizophrenia, schizophreniform, schizoaffective, delusional disorder, psychotic disorder not otherwise specified, and brief psychotic disorder according to the Diagnostic and Statistical Manual of Mental Disorders, Fifth Edition (DSM-V). The criteria for affective psychotic disorders is the DSM-V diagnosis of major depressive disorder with psychosis or bipolar disorder with psychosis (38). Please see Table S1., (38) and https://www.humanconnectome.org/storage/app/media/documentation/HCP-EP1.1/HCP-EP_Release_1.1_Manual.pdf for more details.

#### 2.1.2. Image acquisition and processing

Imaging data were collected on three Siemens MAGNETOM Prisma 3T scanners, with 32-channel head coils at Indiana University and Brigham and Women’s Hospital, and a 64-channel head and neck coil at McLean, but the neck channels were turned off (https://www.humanconnectome.org/storage/app/media/documentation/HCP-EP1.1/HCP-EP_Release_1.1_Manual.pdf). Special procedures and analyses had been taken to ensure the homogeneity of the image quality across both sites (38). The Connectome Coordinating Facility at Washington University provides image processing, central quality control, and data coordination services for all the HCP-style datasets (39). The HCP-EP datasets underwent the same protocol as the HCP lifespan dataset (39). In brief, the T1-weighted structural MRI scans applied a multi-echo MPRAGE sequence with a high isotropic resolution (0.8 mm; other parameters include T1 1000 ms, TR 2400 ms, and 208 slices; see (39) and https://www.humanconnectome.org/study/hcp-lifespan-aging/project-protocol/imaging-protocols-hcp-aging for details). The HCP-EP dataset included cortical surface meshes created by the FreeSurfer-HCP pipeline (40–43), which is based on FreeSurfer version 6.0 (41) with HCP-specific enhancements (40), from T1-weighted MRI images. The cortical surface mesh was further downsampled and registered on the fsLR-32k template, with 32,492 vertices on each hemisphere of the cortex (44). The fsLR-32k template provides an accurate cortical shape of the standard Montreal Neurological Institute (MNI) template (40) but is less computational demanding than the native MNI surface mesh model, and thus the space of the fsLR-32k template is recommended when analyzing the data at the vertex level (40). We used the registered images provided by the HCP-EP dataset without further corrections or smoothing.

#### 2.1.3. Clinical and medication assessment

The HCP-EP dataset provided the severity of psychotic symptoms of EP patients measured by the Positive and Negative Syndrome Scale (PANSS (45)). We followed van der Gaag et al. (46) to employ a five-factor model that has been widely used for evaluating psychotic symptoms (47–52). The five-factor model is more robust and clinically relevant than the original three-factor model (46, 50, 52). The five-factor model comprises the following dimensions: positive symptoms, negative symptoms, disorganization symptoms, excitement, and emotional distress, and was constructed by ten-fold cross-validation with more than 5000 subjects (46). We used factor analysis with maximum likelihood estimation and oblique rotation (53, 54) as implemented in (55, 56) to extract the five factors from the items that had occurred across all folds of the cross-validation in van der Gaag et al. (46). To confirm the robustness of our results, we also tested the five-factor model using the items selected by a recent meta-analysis (57), yielding similar results (please see Table S2 for the items of each factor).

HCP datasets provided lifetime exposure duration of antipsychotic drugs and CPZ of current antipsychotic drugs. Both measures were uncorrelated with the MAS eigen-groups in EP patients (*P_FDR_* > 0.8 and uncorrected *P*-values all >0.5), suggesting that medication has a limited influence on cortical shape asymmetries.

### 2.2. Spectral shape analysis

To obtain a multiscale shape description of the left and right cortical surface meshes obtained from FreeSurfer, we solved the eigenfunction-eigenvalue problem of the Laplace-Beltrami operator (LBO), which is given by (33–35):

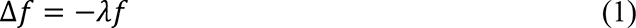

where Δ is the LBO and *f* is a distinct eigenfunction with corresponding eigenvalue }. . The eigenfunctions are orthogonal and describe shape variations at different spatial wavelengths, ordered from coarse-grained (the second eigenfunction in Fig. 1A) to fine-grained scales (e.g., the 500^th^ eigenfunction in Fig. 1A) (34, 35, 58–60). The eigenvalues form a sequence that ranges from zero to infinity, i.e., 0 ≤ }.^l^ ≤ }.^2^≤ …< ∞, and each eigenvalue is analogous to the wave frequency of sinusoids in Fourier analysis (21, 34, 35). The family of eigenfunctions thus represents an orthogonal basis set that can be used to fully decompose and reconstruct the shape of each cortical hemisphere (35). An intuitive example of the LBO eigenvalues and eigenfunctions is percussion a drum: eigenvalues are analogous to the vibration frequency of the drum membrane, and the eigenfunctions are the corresponding vibration patterns of the drum membrane (59).

**Fig. 1.**
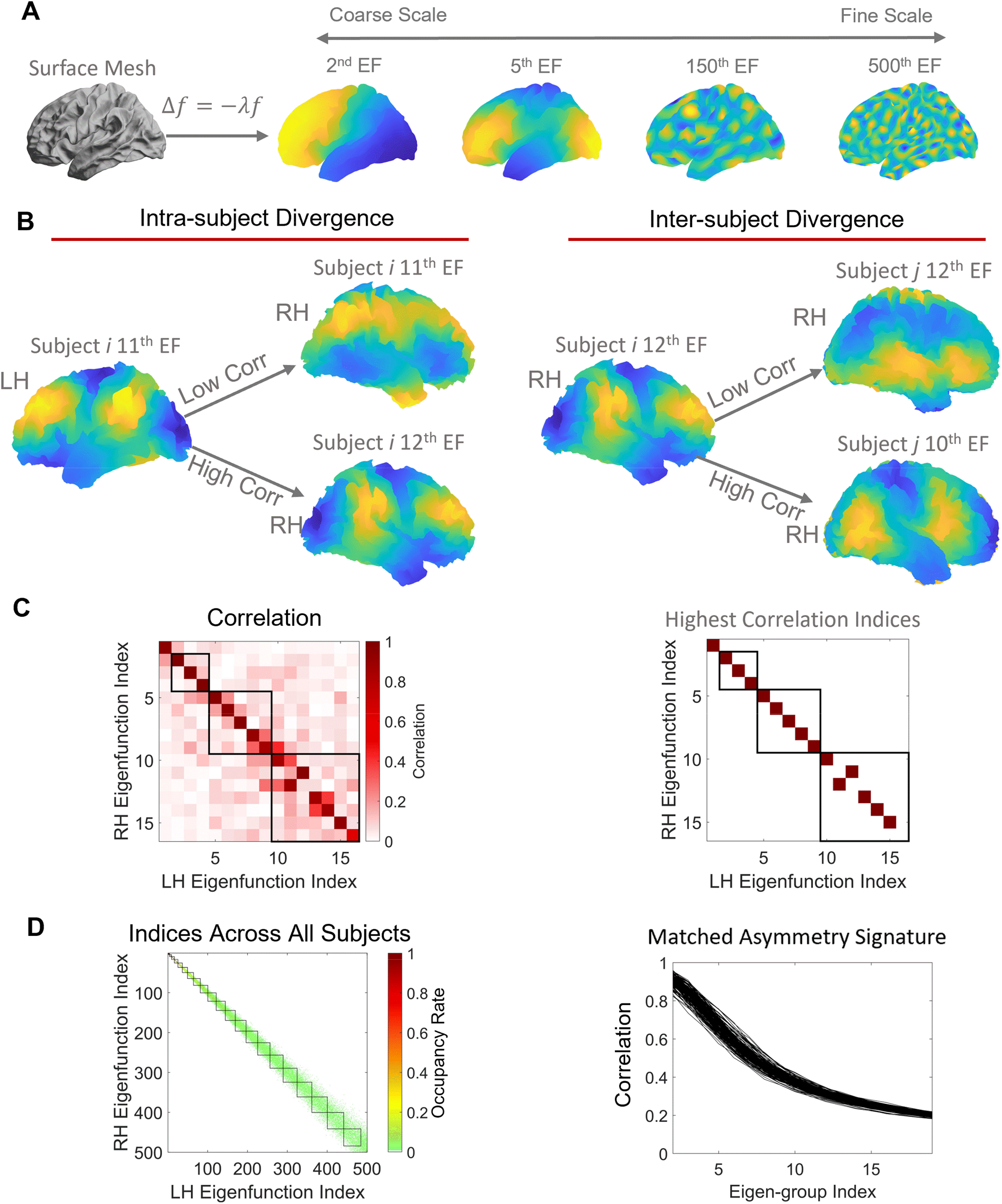
Schematic of the analysis workflow. (**A**) The shapes of the left and right hemispheres (surface mesh) were independently decomposed using the eigenfunctions of the Laplace-Beltrami Operator (LBO). The eigenfunctions describe shape variations at different spatial scales, ordered from coarse (the second eigenfunction, EF) to fine (e.g., the 500^th^ eigenfunction) scales. (**B**) Example of intra- and inter-subject divergences in eigenfunction patterns. The left panel shows that the 11^th^ left eigenfunction is clearly more similar to the same individual’s 12^th^ right eigenfunction than the 11^th^ right eigenfunction. The right panel shows that one individual’s 12^th^ right eigenfunction is more similar to another individual’s 10^th^ right eigenfunction than the 12^th^ right eigenfunction. (**C**) Left panel, correlations between the left and right eigenfunctions from the individual in the left panel of (**B**). From this correlation matrix, we plotted the indices with the highest correlations across all 500 analyzed eigenfunctions in the right panel. The colored squares are the indices with the highest correlations. Here, we only show the first 16 eigenfunctions, and the squares are the boundaries between eigen-groups (see section 2.4*. Cortical shape harmonics* for details). For each row of the matrix, we take the maximum absolute correlation observed. The vector of these correlations across rows is the matched asymmetry signature (MAS). (**D**) Left panel, the mean of the highest indices across all subjects with respect to boundaries between eigen-groups as the squares. Most maximal correlations occurred within the same eigen-groups, but some of them appeared in adjacent eigen-groups. Right panel, we take the mean of MAS across each eigen-group and its adjacent two eigen-groups. This eigen-group based MAS collapses the 500 eigenfunction-specific values to 20 eigen-group-specific values. Here, each line represents one individual. Next, we compare the MAS between the EP patients and the HC and use the eigen-groups with significant group differences to analyze their relationships with psychotic symptoms. Panel (A) was adapted from (21).

We utilized the Shape-DNA software package (33–35) to perform an eigendecomposition of the cortical surface mesh of each hemisphere of an individual (40–43). Shape-DNA uses the cubic finite element method to solve and optimize the eigenfunction/eigenvalue problem of the LBO based on the intrinsic shape of an object or cortex (34, 35). Shape-DNA outperforms other shape analysis methods for retrieving object shapes (61) and has been applied to characterize brain shapes with superior subject identifiability than other conventional measures (21, 33) such as cortical volume, thickness, gyrification, and inter-regional functional coupling (62).

### 2.3. Matched asymmetry signature (MAS)

In our previous work, we directly calculated the differences between the LBO eigenvalue spectra of the left and right hemispheres to characterize the fluctuating asymmetry of cortical shape across a spectrum of spatial scales, also called the shape asymmetry signature (SAS) (21). The SAS is very sensitive for quantifying individualized shape asymmetry features (21) but is too individualized to facilitate group comparisons, as the spatial patterns defined by higher-frequency eigenfunctions are often highly divergent to allow simple pooling across individuals. We therefore developed a new spectral approach for quantifying cortical shape asymmetry that is more suitable for group comparisons and more accurately accounts for hemispheric differences in the spatial patterning of shape variations. Instead of using the eigenvalues, we calculated the product-moment correlation between the left and right eigenfunctions to characterize the divergence of the two hemispheres in each individual.

A critical challenge in this regard is that there is no guarantee that the eigenfunctions of the left and right hemispheres are directly comparable. As shown in Fig. 1B, eigenfunctions of the left and right hemispheres do not necessarily overlap at the same index (i.e., the maximum correlation may occur at different indices); in this particular individual, the 11^th^ eigenfunction of the left cortex is more similar to the 12^th^ eigenfunction (*r* = 0.96) of the right cortex than the 11^th^ right eigenfunction (*r* = 0.17) (Fig. 1B, left panel). This occurs because subtle variations in the shape of the left and right cortices can alter the ordering of eigenfunctions, and sometimes result in quite distinct eigenfunctions at higher spatial frequencies. Although the difference in the ordering of the left and right eigenfunctions reflects individual shape variations, this divergence makes it difficult to compare eigenfunctions across individuals. In this study, after obtaining 500 eigenfunctions for each hemisphere of each individual (Fig. 1A), we first estimated correlations between the spatial pattern of each pair of left-right eigenfunctions, resulting in a symmetric 500 × 500 matrix of correlations quantifying the similarity between left and right eigenfunctions across spatial scales (e.g., Fig. 1C, left panel). The maximum absolute values of correlations observed across the rows of this matrix (or equivalently, columns) quantify the similarity between optimally-matched eigenfunctions of the left and right hemispheres at each of the 500 spatial scales considered in the decomposition (e.g., Fig. 1C, right panel). The resulting vector of 500 maximum correlations thus provides a multiscale, individualized description of cortical shape asymmetries, which we term the Matched Asymmetry Signature (MAS), with lower MAS values corresponding to higher shape asymmetry. We take the absolute value of the correlation because the sign of a given eigenfunction is arbitrary.

### 2.4. Cortical shape harmonics

In the case of a perfect sphere, the eigenvalues and eigenfunctions of the LBO can be sorted into distinct groups, called spherical harmonics. Within the same spherical harmonic group, the eigenvalues are degenerate (i.e., identical) and the corresponding eigenfunctions describe shape variations at similar spatial wavelengths but spatially varying along orthogonal axes (60). There are 2(*L* + 1) – 1 eigenfunctions in the *L*^th^ group (21, 60): the first eigen-group (*L* = 1) is comprised of the 2^nd^ to 4^th^ eigenfunctions, the second eigen-group (*L* = 2) is comprised of the 5^th^ to 9^th^ eigenfunctions, and so on. The first eigen-group describes the shape variations at the coarest spatial scales, which are the variations along the *X, Y, Z* axis. Higher eigen-group measures shape variatios at finer spatial scales (please see Fig 2B for the spatial scales of the eigen-groups). It has been shown that these spherical harmonic groups are roughly conserved for the cortex, since the cortex is topologically equivalent to a sphere (60). We can therefore use an approximation of the spatial wavelength in the spherical case to estimate the corresponding wavelength of each cortical eigen-group (21):

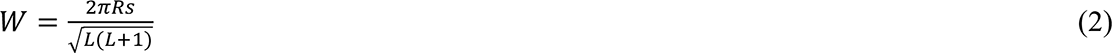

**Fig. 2.**
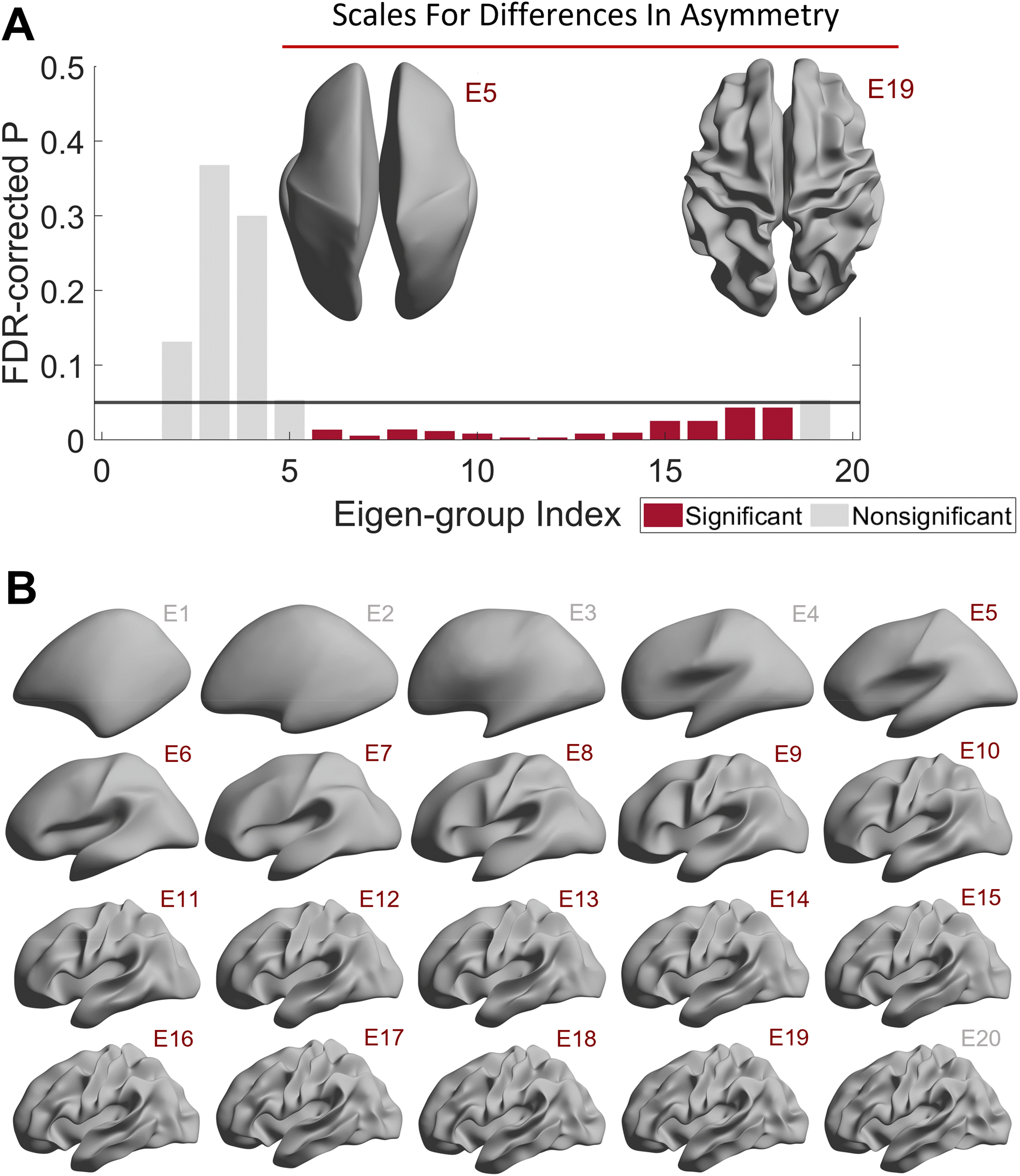
Early psychosis is associated with scale-specific differences in cortical shape asymmetry. **(A)** The 6^th^ to 18^th^ eigen-groups of the MAS are different between HC and EP groups. We used a sliding window approach for MAS, and thus, the wavelength of the 6^th^ to 18^th^ eigen-groups of the MAS are equivalent to the 5^th^ to 19^th^ original eigen-groups, which are between ∼22 (left inset) and ∼75 (right inset) mm. The cortical surfaces are reconstructed from a population-based template (fsaverage). **(B)** Cortical surfaces were reconstructed at different spatial wavelengths from the first eigen-group (E1) and incrementally adding more eigen-groups to the first 20 eigen-groups (E20). Note, these groups are based on the original eigen-groups. Panel (B) was adapted from (21).

where *Rs* is the corresponding sphere radius of the original object (*Rs* is about 67 mm for the population-based template, i.e., fsaverage in FreeSurfer), and *L* is the index of the eigen-group. Accordingly, because the ordering of specific eigenfunctions can vary not only within individuals but also between individuals (e.g., Fig. 1B, right panel), it is more appropriate to focus on the groupings of the eigenfunctions, rather than specific eigenfunctions themselves, when considering scale-specific asymmetries.

Figure 1 C and D indicate that although most of the highest left-right correlations occur for eigenfunctions from the same eigen-group, some of the maximal correlations are found in adjacent groups. For instance, the right panel of Fig. 1C shows that, the maximal correlation between the 11^th^ eigenfunction of the left cortex is the 12^th^ eigenfunction of the right cortex (from the subject in Fig. 1B, left panel), and therefore, the indices of the maximal correlations are not entirely on the diagonal line, but both 11^th^ and 12^th^ eigenfunctions are within the 3^rd^ eigne-group (the third black square, which encompasses the 10^th^ to 16^th^ eigenfunctions). We then took the average of the indices with the maximal correlations across all subjects to serve as occupancy rates in the left panel of Fig. 1D. For example, if the occupancy rate of the 401^st^ left eigenfunction and the 400^th^ right eigenfunction is 0.2, it means that 20% of the subjects’ 401^st^ left eigenfunction correlated the most with the 400^th^ right eigenfunction. Fig. 1D, left panel also shows that the maxiaml left-right correlations generally on or around the diagonal line and occur within the same eigen-group (each black square represents each eigen-group) or its adjacent two eigen-groups.

We therefore used a sliding window approach and took the mean of MAS values within each eigen-group and its two adjacent groups to collapse the MAS from 500 eigenfunction-specific values to 20 group-specific values. For example, we took the mean correlations across the 1^st^ to 3^rd^ eigen-groups (2^nd^ to 16^th^ eigenfunctions) to represent the second eigen-group of MAS and the mean across the 2^nd^ to 4^th^ eigen-groups (5^th^ to 25^th^ eigenfunctions) to represent the third eigen-group. In this study, we ignored the first eigen-group because it represents the coarsest spatial scale (∼170 mm wavelength) and is highly conserved, providing little individual-specific information. We also did not consider the 20^th^ eigen-group of MAS (362^nd^ to 484^th^ eigenfunctions), since the mean MAS across all subjects was low (i.e., *r* < 0.2, SD = 0.008), indicating that very fine-scale variations in the shapes of the left and right hemispheres are largely independent. We excluded data for one individual who showed eigen-group-specific MAS values that were more than three standard deviations below the sample mean.

Previous studies have shown that the shape measures of the LBO are robust to image noise (21, 58). To confirm the robustness of the MAS to image quality, we used the Euler numbers of the FreeSurfer, a widely used approach (21, 63–65) to quantify the image quality. We took the mean of the Euler number from the left and right hemispheres and calculated the then calculated the Pearson’s correlation between the mean Euler number and the eigen-group-specific MAS values. All eigen-groups of the MAS were unrelated to the Euler number (absolute *r* values all < 0.08; *P_FDR_* all > 0.8). The results confirms that image quality generally does not influence the eigen-groups of MAS.

To summarize, we characterize the shape of each cortical hemisphere using the eigenfunctions of LBO, which offers a natural mathematical description of how the shape of an object varies through space. Eigenfunctions of the LBO correspond to fundamental spatial patterns (akin to axes or modes) of shape variation through space that are intrinsic to each cortical hemisphere of each individual. The corresponding eigenvalues correspond to the spatial frequency, or wavelength, of each eigenfunction, and are ordered such that low values corresponding to coarse-scale shape variations (e.g., broad anterior-posterior gradients; see Fig. 1A, 2^nd^ EF) whereas high values correspond to fine-scale shape variations (e.g., local sulcal and gyral architecture; see Fig. 2A, 500^th^ EF). The MAS is estimated as the correlation of the spatial pattern between a given eigenfunction in the left hemisphere and the most similar eigenfunction in the right hemisphere. It thus represents the degree of left-right similarity in fundamental patterns of shape variation. Since the cortex is topologically equivalent to a sphere, and sets of eigenfunctions in the spherical case correspond to shape variations with the same wavelength, we can group eigenfunctions into eigen-groups of spatial patterns with similar wavelengths (see fig 2B). Averaging across Eigen-groups thus provides a more robust index of scale-specific shape asymmetries. Critically, our approach isolated asymmetries in cortical shape as distinct from those in size. This is critical, since shape asymmetries specifically are highly unique to individual brains and are under the influence of unique environmental influences, rather than genetic or common environmental factors (21). They thus offer a window into brain changes related to environmental risk factors.

### 2.5. Size-based anatomical asymmetry

To compare the effects of shape asymmetry to other commonly used morphological asymmetries, including cortical thickness, surface area, and LGI (the ratio between pial surface and outer smoothed surface automatically measured by FreeSurfer (66)), we applied a widely used asymmetry index (16, 67, 68) to quantify these non-shape-based morphological asymmetries as:

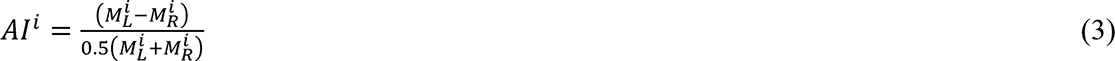

where Al^i^is the asymmetry index of subject *i*, 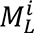 is the mean value (mean of the whole hemisphere or region) of the morphological measurement, i.e., cortical thickness, surface area, or LGI, from subject *i*’s left hemisphere, and 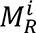 is the value from the right hemisphere. We registered each subject image on the fs_LR 32k template (44) and calculated the asymmetry at both global (i.e., whole hemisphere) and regional levels. For the regional level, we calculated the cortical thickness, surface area, and LGI of each region defined in the Desikan-Killiany (69) or HCPMMP1 (70) atlases. There are 35 and 180 regions in each hemisphere of the Desikan-Killiany and HCPMMP1 atlases, respectively, but we excluded the hippocampus from the HCPMMP1 atlas as cortical thickness cannot be measured for this structure.

### 2.6. Statistical analysis

After calculating the eigenfunctions of LBO from shape-DNA algorithm (34, 35), the resulting eigenfunctions were further analyzed by using the MATLAB (version R2020b). We used a general linear model (GLM) to analyze the differences of each eigen-group between HCs and EP patients as well as between EP patients with non-affective and those with affective psychosis, controlling for sex, age, neuroimaging sites, and total brain size as confounding variables. We did not control for the medication effects because both exposure duration of antipsychotic drugs and CPZ of current antipsychotic drugs were unrelated to all the eigen-groups considered in this study (*P_FDR_* > 0.8) in EP patients. We also did not control for handedness because previous studies have found that cortical shape asymmetry is unrelated to handedness (21, 33). Eigen-groups analyzed in this study include their adjacent groups and are not independent of each other, and thus, we used a permutation test with 50,000 iterations for statistical inference. We used a tail approximation implemented in comparing the original GLM coefficients with permuted coefficients to ensure reliable *P*-value estimations (71) and controlled the false discovery rate (FDR, *q* = 0.05) to correct for multiple comparisons.

We examined correlations between the eigen-groups of MAS and PANSS factors using canonical correlation analysis (CCA) on the 104 EP patients who responded to all items of the PANSS (45). We first used principal component analysis (PCA) to reduce dimensionality and minimize collinearity of the eigen-groups of MAS at different scales. We then applied the CCA to identify maximal covariance (72) between the PCs of MAS eigen-groups and PANSS factors by linear combinations, controlling for age, sex, neuroimaging sites, and total brain size as confounding variables. The *P*-values of the canonical modes were calculated using a recently-developed CCA permutation procedure (72) with 50,000 iterations and controlling family-wise error rate (FWER) (72). We used bootstrapping with 1000 iterations to identify reliable estimates of loadings of each PANSS factor on the canonical variate (21, 73), and the resulting *P*-values were then corrected for multiple comparisons by FDR (*q* = 0.05). We also applied bootstrapping with 1000 iterations and FDR correction to measure the reliable correlations between each eigen-group of the MAS and the canonical variate.

## 3. Results

### 3.1. Increased cortical shape asymmetries at coarse scales in EP patients

We first used separate GLMs to compare the 2^nd^ to 19^th^ eigen-groups of MAS between patients and controls. We found that the 6^th^ to 18^th^ eigen-groups (Fig. 2A), spanning wavelengths between about 22 and 75 mm, were significantly different between the HC and EP groups (*P_FDR_* < 0.05; Cohen’s *d* = 0.28−0.51; Fig. S1), with the MAS in the EP group being significantly lower than the HC group. In other words, patients showed greater asymmetry in shape than controls. The distributions of the MAS of these two groups are shown in Fig. S1. Although the 6^th^ to 18^th^ eigen-groups were all significantly different between the HC and EP groups, the effect sizes of some eigen-groups, such as the 11^th^ and 12^th^ eigen-groups (*d* = 0.51 and 0.49, respectively), were higher than some other eigen-groups, such as the 17^th^ and 18^th^ eigen-groups (*d* = 0.29 and 0.28, respectively) (Fig. S1). In contrast, we found that commonly used morphological asymmetries, i.e., based on cortical thickness, surface area, and LGI at both global (GLM *P*-values > 0.05; Fig. S4) and regional levels using either the HCPMMP1 or Desikan-Killiany atlases, were not different between these two groups (GLM *P_FDR_* > 0.05; Table S3 and S4). Thus, asymmetries of cortical shape are more sensitive than the traditional morphological measures in discriminating HC and EP individuals.

Next, we compared patients with affective psychosis and those with non-affective psychosis. All eigen-groups (2^nd^ to 19^th^) of the MAS were not significantly different between groups (*P_FDR_* > 0.05).

### 3.2. Cortical shape asymmetries correlate with symptom severity

We used CCA (72) to examine relationships between the eigen-groups of MAS and the five factors of psychotic symptoms measured by the PANSS. We applied PCA to the 6^th^ to 18^th^ eigen-groups, which showed differences in our group analysis (see Fig. 2), and retained the first two principal components (PCs), which explained 92% of the variance.

The CCA identified a single canonical mode that was statistically significant (CCA *r* = 0.45; *P_FWER_* = 0.002; Fig. 3A). The loadings of the first two PCs (0.54 and 0.83) of the MAS on the canonical variate were all significant (*P_FDR_* < 0.0001), and the 6^th^ to 9^th^ eigen-groups were positively correlated with the canonical variate (*P_FDR_* < 0.05; Fig. 3B), with corresponding wavelength range of ∼40 to ∼75 mm (Fig. 3B insets). The loading of PANSS factors was negative (*P_FDR_* < 0.001) for excitement (representing impulsivity (50)) and positive (*P_FDR_* < 0.001) for emotional distress (Fig. 3C), indicating that patients with higher cortical shape asymmetry show more impulsivity symptoms and less severe emotional distress. We also confirmed that the antipsychotic drug exposure time was not correlated to all PANSS factors (*P_FDR_* > 0.2).

**Fig. 3.**
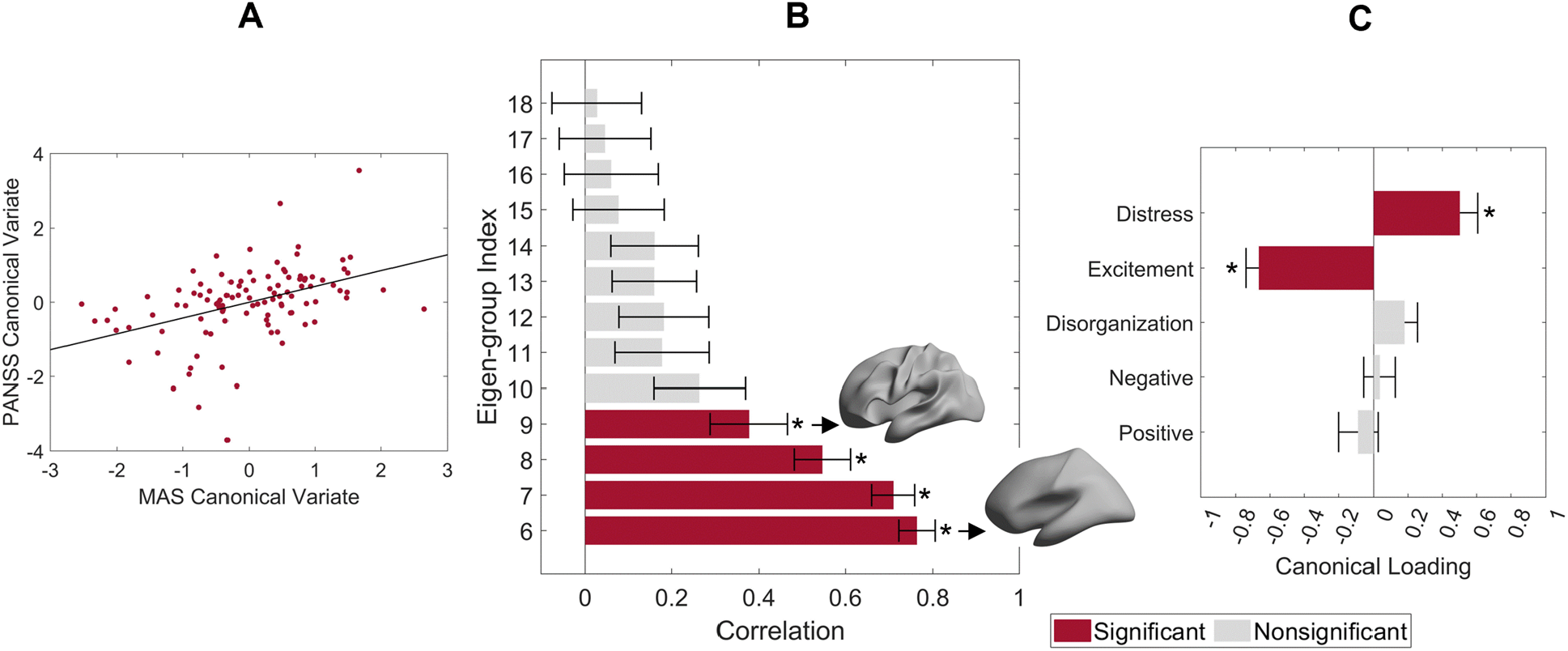
Individual differences in the matched asymmetry signature (MAS) are correlated with symptom severity. **(A)** Association between the canonical variates of the first two PCs of the eigen-groups of MAS and the five PANSS factors with the corresponding least-squares regression line in black. **(B)** Correlations between the 6^th^ to 18^th^ eigen-groups of the matched asymmetry signature and the corresponding canonical variate; the 6^th^ to 9^th^ eigen-groups are significant. Insets represent the range of the corresponding spatial wavelengths. **(C)** Canonical variate loadings of the PANSS factors; the excitement and emotional distress factors are significant. For panels B and C, the error bars represent ± 2 bootstrapped standard errors (SE), and the asterisks denote bootstrapped *P_FDR_* < 0.05.

## 4. Discussion

Altered brain asymmetries have frequently been reported in patients with psychosis. Here, we applied a multiscale approach to examine abnormalities of cortical shape asymmetry, independent of variations in regional size, in EP patients. We found that EP patients showed a higher degree of cortical shape asymmetry across a range of coarse spatial scales when compared to HC. In contrast, no group differences in asymmetries in cortical thickness, surface area, and LGI were identified.

EP patients with a higher cortical shape asymmetry showed more severe excitement and less severe emotional distress. Together, these findings support the sensitivity of our spectral approach for characterizing shape asymmetries in psychosis, indicating that altered asymmetries of shape are expressed at certain spatial wavelengths, and that individual differences in these asymmetries are related to symptom severity.

### 4.1. EP patients show increased asymmetry of cortical shape at coarse scales

Surprisingly, we found that EP patients had greater left-right asymmetry of cortical shape when compared to controls. This finding contradicts to many previous studies reporting reduced asymmetries in patients (19, 20, 29, 30, 36). Conventional asymmetry studies use size-related measures, such as volume or cortical thickness, which can conflate variations in shape and size (21). The distinction between shape and size-based features is crucial because two objects can have similar volumes but with very different shapes (34, 74). Although the literature on shape asymmetry in patients with psychosis is very limited, one prior study has also found an increased asymmetry of global cortical shape in patients with schizophrenia (28). Núñez et al. (28) applied dice coefficient to measure cortical asymmetry by calculating the ratio of the intersection area between the original cortex (e.g., right cortex) and flipped cortex (new right cortex that was flipped from the left) to the total area of the original cortex and the flipped cortex. However, this approach cannot disentangle the effects of shape form size and cannot measure scale-specific effect. For an extreme example, even if two hemispheres are symmetric in shape but one hemisphere is proportional larger than the other, the dice coefficient may still be the same as another subject whose two hemispheres are asymmetric in shape but identical in volume. Previous study has shown that isolating the shape asymmetry from the size effect providing superior subject identifiability. Unlike dice coefficient, the MAS disentangles the shape asymmetry from the size effects and decomposes the asymmetry at different spatial scales. Our spectral approach suggests that this increased asymmetry is apparent within a specific spatial wavelength range between ∼22 mm and ∼75 mm. Among this range, the strongest effect size occurred at the spatial wavelength of about 37 mm, while the effect sizes decrease at finer spatial wavelength. The results are in line with our previous study (21) that found the optimal subject identifiability of cortical shape asymmetry occurred at the spatial wavelength of about 37 mm.

Our scale-specific approach is akin to analyzing the seismic wave frequencies of earthquakes at the global tectonic level, whereas classical point-wise approaches are analogous to only focusing on a specific city (21), which may miss broader patterns. Indeed, grey matter abnormalities in patients with psychosis are often widespread (13, 75). However, such deviations may show minimal overlap in specific brain locations across patients (76–78), potentially reflecting the known clinical heterogeneity of the condition (76, 79). The vast majority of past work on cerebral asymmetries in psychosis has focused on size-related measures, estimated at global, regional, or point-wise levels (13, 20, 29, 36), and are insensitive in capturing diffused and heterogeneous brain abnormalities in patients with psychosis. Our analysis found no differences in the asymmetry of size-related measures at global or regional levels, which aligns with other works showing limited evidence for altered asymmetry of these measures in people with psychosis (11–14, 30). Indeed, the findings have been highly inconsistent, with one meta-analysis showing that even for the superior temporal gyrus −the region most widely reported as showing abnormal asymmetry in patients with schizophrenia− about half of the studies do not find evidence of significantly altered size-based asymmetry (10). Similarly, although some studies have reported decreased LGI values in patients with psychosis or those risk groups (80–82), the results were not consistently reproduced, with some studies showed increased gyrification or absent of the effects (30, 83–85). In this study, we found no differences in the LGI asymmetry at both global or regional levels. Our results were in line with previous study that shows intrinsic cortical shapes derived from the LBO are superior than the LGI in identifying individual variations (33). Although LGI is related to the cortical shape, it measures the gyrification at vertex level, and the result of no group effects on LGI asymmetry is in line with our MAS finding that the asymmetry effects are shown at coarse scales. Moreover, similar to the size-related measures, LGI is also a point-wise approach and is insensitive to the diffused abnormalities. Our findings suggest that the spectral approach developed here offers a new window in understanding asymmetries of cortical shape. The MAS measures the cortex as a whole but characterizes the shape variations of it across multiple spatial scales and is more sensitive to psychosis-related brain changes than other traditional morphological measures, such as cortical thickness, surface area, or LGI. Replication of these findings in independent samples will be an important extension of this work.

Our spectral approach offers a natural way of characterizing fluctuating asymmetries, which capture individual-specific brain phenotypes (21). Fluctuating asymmetries are hypothesized to arise from developmental instability or individual-specific genetic perturbations (27, 28), but it has been shown that fluctuating asymmetries in cortical shape are mainly driven by person-specific environmental influences (21). Many studies have suggested that environmental factors in early life, such as maternal stress, infections, nutrition during pregnancy, childhood adversity, and stress, may affect neurodevelopment and contribute to psychosis (7, 86). Studies of fluctuating asymmetry in the human brain are limited, but some have found that fluctuating asymmetries of the left-right sides of the human body, such as asymmetry of left and right fingerprints, are also increased in patients with psychosis (26, 87–89).

### 4.2. Cortical shape asymmetry is correlated with psychotic symptoms

EP patients with higher cortical shape asymmetries at coarse scales showed more severe excitement symptoms and less severe emotional distress. These coarse spatial scales correspond to wavelengths of about 40 to 75 mm. Notably, this range is within the spatial scales previously found to be optimal (larger than 37 mm) for distinguishing individual brains and related to general cognitive functions (21). Together, these findings suggest that cortical shape asymmetries at coarse scales, but not fine scales, contain personalized brain features, and these features have implications for both normal and abnormal brain function. Past studies using point-wise approaches have found associations between cortical asymmetries of size-based measures and hallucinations (37, 90), but the findings have not been consistently replicated. For example, Ohi et al. (30) did not find any correlation between the asymmetries of volume, thickness, and surface area in the superior temporal gyrus with any clinical symptoms.

The excitement factor of the PANSS may be more related to ADHD and oppositional defiant disorder, while the emotional distress factor is related to anxiety, depression, and stress. Studies have found abnormalities in cortical asymmetry among the above disorders (7, 91, 92) and have suggested that various forms of stress are key risk factors of many mental disorders and may also influence normal brain development, resulting in abnormal brain asymmetries (7). However, we did not find differences in MAS between patients with affective psychosis and those with non-affective psychosis. This result may be due to the small sample size of patients with affective psychosis (n = 32).

## 5. Limitations and Future Directions

In this study, we analyzed EP patients, which minimizes the effects of prolonged medication. Nonetheless, about 78% of the patients had prior exposure to antipsychotics. Although we found that antipsychotic drug exposure time and current CPZ value were unrelated to MAS, the long-term effects of medication on shape asymmetries remain unclear. Future work should aim to replicate our results on medication-naive patients with first-episode psychosis or to compare the results in different psychosis stages, such as chronic schizophrenia. Moreover, it would also be beneficial for future studies to utilize this novel methods to a wide range of neuropsychiatric and neurological diseases, such as ADHD, autism, bipolar disorders, and dementia, given that abnormal brain asymmetries have been reported in these diseases (7, 17, 93, 94). Finally, the MAS also has the potential to measure the development of cortical asymmetry in children and adolescents or its relationships with brain age.

## 6. Conclusion

We developed a novel method to derive a multiscale characterization of cortical shape asymmetries and showed that patients with early psychosis displayed increased asymmetries at coarse scales. In contrast, asymmetries of cortical thickness, surface area, and gyrification were not different between patient and control groups. We also found that patients with a higher degree of cortical shape asymmetries at coarse scales showed more severe excitement symptoms and less severe emotional distress. Together, these findings indicate that cortical shape asymmetries are more sensitive than other morphological asymmetries to capturing differences between patients with psychosis and healthy controls, and these asymmetry features were related to the excitement and emotional distress symptoms.

## Data Availability

The data is open source data. Please refer to https://www.humanconnectome.org/study/human-connectome-project-for-early-psychosis

## 7. Acknowledgments

A.F. was supported by the Sylvia and Charles Viertel Foundation, National Health and Medical Research Council (IDs: 1197431 and 1146292), and Australian Research Council (ID: DP200103509). J.T. was supported by a Turner Impact Fellowship from the Turner Institute for Brain and Mental Health, Monash University. Data were provided by HCP-EP: Principal Investigators: Alan Breier, Martha Shenton.

## 8. Declarations of interest

K.M.A. is a scientific advisor and shareholder of BrainKey Inc., a medical image analysis software company. The other authors declare that they have no competing interests.

## 9. Ethics Statement

All participants provided their written informed consent to participate in the HCP-EP, and the HCP-EP was reviewed and approved by the Human Connectome Project. The HCP-EP complied with the ethical standards of the relevant national and institutional committees on human experimentation and with the code of ethics of the World Medical Association (the Helsinki Declaration of 1975, as revised in 2013).

## Notes

### Competing Interest Statement

A author (K.M.A) is a scientific advisor and shareholder in BrainKey Inc, a medical image analysis software company

### Author Declarations

We use HCP for Early Psychosis dataset, which is a open source data and opened before the start of our study. All details and locations of the data, please refer to: https://www.humanconnectome.org/study/human-connectome-project-for-early-psychosis

